# Joint Temporal Patterns By Integrating Diet and Physical Activity

**DOI:** 10.1101/2023.01.23.23284780

**Authors:** Jiaqi Guo, Luotao Lin, Marah M. Aqeel, Saul B. Gelfand, Heather A. Eicher-Miller, Anindya Bhadra, Erin Hennessy, Elizabeth A. Richards, Edward J. Delp

## Abstract

Both diet and physical activity are associated with obesity and chronic diseases such as diabetes and metabolic syndrome. Early efforts in connecting dietary and physical activity behaviors to generate patterns rarely considered the use of time. In this paper, we propose a distance-based cluster analysis approach to find joint temporal diet and physical activity patterns among U.S. adults ages 20-65. Dynamic Time Warping (DTW) generalized to multi-dimensions is combined with commonly used clustering methods to generate unbiased partitioning of the National Health and Nutrition Examination Survey 2003-2006 (NHANES) dataset. The clustering results are evaluated using visualization of the clusters, the Silhouette Index, and the associations between clusters and health status indicators based on multivariate regression models. Our experiments indicate that the integration of diet, physical activity, and time has the potential to discover joint temporal patterns with association to health.

## I. Introduction

Diet and physical activity are known to be associated with obesity and chronic diseases [21], [24], [34], [40], [56]. Both dietary and physical activity behaviors occur in a sequential manner, following certain rhythms that start and end throughout the day [60]. There is a growing interest in the temporality of diet and physical activity (e.g. time of eating and exercise) and their effects on health. A few studies have explored the use of cluster analysis approaches to find patterns of diet or physical activity independently while integrating the time of eating or exercise behaviors [14], [15], [22], [23]. These patterns derived with temporal information were proven to be associated with obesity and chronic diseases [11], [37], [53]. These studies demonstrated the advantages of using data-driven methods for the integration of time in diet/physical activity pattern analysis. In [30], [44], [58], the authors showed that dietary and physical activity behaviors could be connected in complex ways and have a synergistic impact on health. Some early efforts have been made to investigate the connections between dietary and physical activity behaviors and generate joint diet and physical activity patterns [54], [58], [61]. Less is known about the interaction of time and diet and physical activity and their joint influences on health.

In this paper, we describe the Joint Temporal Diet and Physical Activity Pattern (JTDPAP). This is defined as the partitioning of a joint diet and physical activity dataset using data-driven methods (such as cluster analysis) that integrate the time of diet and physical activity behaviors. Participants are separated into mutually exclusive clusters based on similar dietary and physical activity characteristics. Each cluster represents a specific joint temporal pattern. We focus on distance-based cluster analysis for finding joint temporal patterns among U.S. adults ages 20-65. The goal of our study is to find joint temporal patterns that have distinguishable diet and physical activity characteristics (e.g., intensity, duration, and timing), and could also be meaningfully connected to health.

To date, the National Health and Nutrition Examination Survey 2003-2006 (NHANES) [5] is one of the only publicly-available, nationally representative datasets that capture dietary intake through 24-hour recall methodology and physical activity through accelerometry devices. The joint diet and physical activity dataset used in this paper is constructed from the NHANES dataset. We use the data from 1836 participants age 20-65 in the NHANES for our analysis. For the physical activity, the NHANES used uni-axial accelerometers to measure minute-level activity intensity in units known as “Physical Activity Count (PAC)” [5]. We randomly select one valid weekday for each participant to form the physical activity dataset in this paper. The physical activity data are one-dimensional time series of length 1440 samples (60 minutes × 24 hours). For the diet information, the NHANES collected the amount and time of energy intakes from two 24-hour dietary recall. We select one weekday to generate the one-dimensional diet time series for the participants. Each participant in our dataset is thus represented by two time series, one for diet and another one for physical activity. We combine the two time series from the diet and the physical activity to form the joint dataset we used in our experiments. Note that the two time series are not temporally aligned in that the diet intakes and physical activities occurred at different days and different times. Our goal is to cluster this joint dataset and evaluate the clusters’ association to health.

In this paper, we explore a distance-based cluster analysis approach. Since both diet and physical activity time series are subject to potential warpings (e.g, having breakfast a few minutes earlier than usual; jogging for 10 minutes longer), we follow our previous studies of temporal diet/physical activity patterns [22]–[24], and choose the Dynamic Time Warping (DTW) [50] as the distance measure to align the samples of input time series. Since DTW was originally designed for comparing one-dimensional time series, we adapt two commonly used methods to generalize DTW to multi-dimensions, namely the independent and dependent multi-dimensional DTW (denoted as *DTW*_*I*_ and *DTW*_*D*_) [52]. In both *DTW*_*I*_ and *DTW*_*D*_, we introduce a parameter *α* ∈ [0, 1] to control the emphasis on physical activity and diet. With larger *α*, the physical activity difference between two participants would have larger impact on their DTW distance. When *α* = 1, both *CDTW*_*I*_ and *CDTW*_*D*_ are only using the physical activity time series. This is described in more detail in Section II-B.

In this paper, three distance-based clustering methods which are commonly used in physical activity or diet pattern research are explored, namely kernel k-means (KKM) [19], [47], spectral clustering (SPEC) [41], and kernel hierarchical agglomerative clustering (KHAC) [31], [45]. To combine the independent and dependent multi-dimensional DTW distances with the clustering methods, the Gaussian Dynamic Time Warping Kernel [12] is used to convert *DTW*_*I*_ and *DTW*_*D*_ into kernel functions. The clustering results are evaluated in a similar way as our previous diet/physical activity studies [22]–[24]. Three criteria are used in the evaluation process, including 1) visualization tool to illustrate the characteristics of dietary and physical activity behaviors of the clusters; 2) internal criteria (the Silhouette Index [46]) to determine the number of clusters; 3) external criteria based on multivariate linear regression (MLR) and multiple comparison to find the clusters’ association to health status indicators.

Our major contributions in this paper are summarized below:

- We extend our previous research on temporal patterns through jointly clustering of diet and physical activity.
- Two ways of generalizing DTW to multi-dimensional time series are described that allow the emphasis on physical activity or diet.
- We evaluate the clustering results through visualizations of the clusters, internal criteria, and external criteria.
- Our experiments show that the integration of diet, physical activity, and time has the potential to find joint temporal patterns with association to health.

## II. Related Work

### A. Joint Diet and Physical Activity Pattern

Here we review and summarize previous work on joint diet and physical activity pattern analysis [14], [15], [18], [37], [39], [49], [51], [53]. We review the work from three aspects: type of data used, clustering method, and evaluation criteria. Boone-Heinonen et al. [14] studied obesity-related diet and physical activity behaviors in an adolescent population. Both diet and physical activity data were collected through survey questionnaires. For diet, 11 composite variables were used to represent the consumption and types of food/beverage consumed. For physical activity, 25 variables that comprised the numbers of weekly instances of different types of activities and the hours of sedentary behavior were collected from the questionnaires. The combined 36 variables (11 diet and 25 physical activity) were clustered using SAS FASTCLUS (Software SAS version 9, Research Triangle Institute, Research Triangle Park, NC, 2004 [14]), which essentially uses the Euclidean distance for measuring the distance between 36 variables and the k-means for the clustering. The final clustering was based on a series of internal criteria including distinctiveness, robustness, and strength of behaviors. Cameron et al. [15] examined the patterns of diet and activity to find how they related to obesity in children and their mothers. The diet data, which was collected through questionnaires, consists of two variables: amount of healthy and unhealthy food consumption. The Mother’s physical activity data was collected using questionnaires, while the children were objectively assessed using uni-axial accelerometers. Both mother and children physical activity data was later converted into two variables: total activity time and total sedentary time in a week. The diet and activity was clustered using hierarchical agglomerative with Ward’s linkage. The authors reported a result for five clusters as “most able to define specific groups of both mothers and children,” but no quantitative criteria was given. Matias et al. [37] used cluster analysis to estimate joint patterns of diet, physical activity, and sedentary behavior among adolescents, and to find their associations with sociodemographic variables. Diet information was collected through seven questions, and summarized into two variables: the number of days in a week eating a healthy or unhealthy diet. Physical activity and sedentary behavior are also assessed through questionnaires, and summarized as the number of days in a week practicing exercises and the number of hours in a regular day being sedentary (e.g., watching television or playing video games). The clusters were derived using the TwoStep Cluster Analysis in SPSS (version 23 SPSS Inc.; Chicago, IL, USA [1]), and evaluated by Bayesian Information Criterion (BIC) and the Silhouette Index [46]. Skalamera et al. [53] used Latent Class Analysis (LCA) to study the association between educational attainment and health-related behaviors. Eight health-related features including binge drinking, regularly participating in physical activity, eating at fast food restaurant often were dichotomized. The LCA was used to group the participants into 3 clusters based on the dichotomized features. Four internal criteria, including log-likelihood, BIC, sample-size-adjusted BIC (ABIC), and Lo-Mendell Rubin (LMR) adjusted likelihood ratio, were used to select the number of clusters and evaluate the clustering results.

From the above discussion, the previous studies on joint diet and physical activity pattern have a wide variety in terms of clustering methods and data [18], [39], [49], [51]. However, the majority of these studies simplified the complex diet and physical activity behaviors into features of intensity, frequency, or duration. The temporal information such as the time of eating and activity was often omitted.

### B. Dynamic Time Warping for Multi-Dimensional Time Series

We choose the Dynamic Time Warping (DTW) [50] as the distance measure to address the temporal misalignment of the input time series. The DTW distance was originally designed for one-dimensional speech signals [50]. In this paper, the participants are represented by a two-dimensional diet and physical activity time series. Therefore, we need a way to generalize DTW for multi-dimensional time series.

As discussed by Shokoohi-Yekta et al. [52], there are two directions to generalize DTW for multi-dimensional time series. The first direction is the dependent multi-dimensional DTW (denoted as *DTW*_*D*_). In *DTW*_*D*_, different dimensions of the input time series are considered to be dependent (or tightly coupled as described in [52]), and *DTW*_*D*_ finds the same alignments for all dimensions of the input time series. Dynamic programming used in DTW is also used to find the alignments for *DTW*_*D*_ [9], [17], [25]. The second direction is the independent multi-dimensional DTW (denoted as *DTW*_*I*_). As the name suggests, *DTW*_*I*_ takes each dimension as an independent one-dimensional time series, and computes the distance for each single dimension using DTW. The *DTW*_*I*_ distance is a weighted sum of the independent DTW distances. Compared to *DTW*_*D*_, *DTW*_*I*_ finds its own alignments for each dimension, and thus has more flexibility for time series whose different dimensions are loosely coupled [13], [38], [55].

In this paper, both *DTW*_*D*_ and *DTW*_*I*_ are explored for the joint diet and physical activity time series. We introduce a parameter *α* ∈ [0, 1] in both *DTW*_*D*_ and *DTW*_*I*_ to control the emphasis on physical activity and diet. With larger *α*, the physical activity difference between two participants would have more impact on their *DTW*_*I*_ /*DTW*_*D*_ distance. When *α* = 1, both *DTW*_*I*_ and *DTW*_*D*_ are computed only using the physical activity time series, and are the same as one-dimensional DTW distance. Analogously, when *α* = 0, *DTW*_*I*_ and *DTW*_*D*_ are the same as one-dimensional DTW computed only using the diet time series.

## III. Proposed Approach

### A. Definitions and Symbols

Let **X** = [**x**_1_, **x**_2_, · · ·, **x**_*M*_] and **Y** = [**y**_1_, **y**_2_, · · ·, **y**_*M*_] be two time series of the same length *M* (*M* ≥ 1). Let **X**[*k*: *l*] = [**x**_*k*_, **x**_*k*+1_, · · ·, **x**_*l*_] be the sub time series of **X** (1 ≤*k* ≤ *l ≤ M*). Time series **X** and **Y** can be either one- or multi-dimensional. We assume that **x**_*i*_, **y**_*j*_ ∈ ℝ^*D*^ with *D* ≥ 1. Let **X**^(*d*)^ be the *d*^*th*^ dimension of **X** where 1 ≤*d* ≤*D*. Then **X**^(*d*)^ is a one-dimensional time series, and the *i*^*th*^ sample of **X**^(*d*)^ 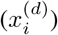 is the *d*^*th*^ element of **x**_*i*_.

### B. One-Dimensional Dynamic Time Warping

Dynamic Time Warping [50] was originally designed for one-dimensional time series such as the marginal diet or physical activity time series. Here we briefly review the basics of DTW before generalizing DTW for multi-dimensional time series. We assume time series **X** and **Y** are both one-dimensional of length *M* in the following discussion.

To compute the distance between time series **X** and **Y**, the Euclidean distance aligns the samples with the same indices and computes the sum of the square differences, i.e.,

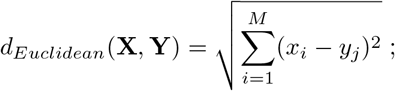

The Euclidean distance is easily affected by small shifts in time, which makes it unsuitable for comparing two time series. In contrast, Dynamic Time Warping provides a way of temporally aligning two time series such that the distance measure is not sensitive to temporal misalignment. The DTW distance between time series **X** and **Y** can be defined as

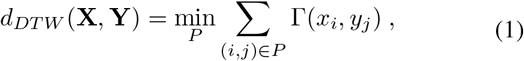

where Γ : ℝ × ℝ → ℝ^+^ is a local distance function which compares a pair of samples. *P* is the warping path which defines how the samples of **X** are aligned to the samples of **Y**. *P* is a contiguous set of index pairs and its element (*i, j*) indicates that the *i*^*th*^ sample of **X** is aligned to the *j*^*th*^ sample of **Y**. In previous temporal pattern studies, the Sakoe-Chiba Band [50] was incorporated in the warping path of DTW as a global constraint. The Sakoe-Chiba Band limits the maximum index difference between aligned samples *x*_*i*_ and *y*_*j*_, i.e., it enforces |*i* − *j*| ≤*T* where *T* is the Sakoe-Chiba Bandwidth. An important reason for introducing global constraints when generating temporal patterns is to prevent potential pathological warpings (e.g., aligning eating events in the morning to eating events in the evening). In this paper, we denote Constrained DTW with the Sakoe-Chiba Band as CDTW.

For detailed discussion of DTW and alternative elastic distances, we refer the reader to the paper by Sakoe et al. [50] and the paper by Marteau [36]. For the use of DTW for independent diet/physical activity pattern analysis, we refer the reader to our previous temporal pattern studies [11], [22]–[24].

### C. Generalize DTW to Higher Dimensions

The definition of DTW in Equation 1 is designed for one-dimensional time series. To apply DTW to the joint diet and physical activity time series, we need a way to generalize DTW to higher dimensions. Previous studies which adapted DTW for multi-dimensional time series [28], [35], [42], [57] can be summarized into two directions: independent and dependent multi-dimensional DTW (*DTW*_*I*_ and *DTW*_*D*_) [52]. In this paper, both *DTW*_*I*_ and *DTW*_*D*_ are explored for the joint diet and physical activity time series.

We consider the input time series **X** and **Y** to be *D*- dimensional (*D >* 1) of the same length *M*. The independent multi-dimensional DTW (*DTW*_*I*_) is defined as the sum of one-dimensional DTW distances computed independently based on each separate dimension. Mathematically, the *DTW*_*I*_ distance between time series **X** and **Y** can be written as:

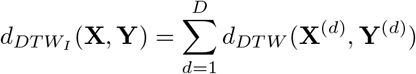

where **X**^(*d*)^ and **Y**^(*d*)^ are the *d*-th dimension of time series **X** and **Y** respectively. *d*_*DTW*_ (**X**^(*d*)^, **Y**^(*d*)^) is the one-dimensional DTW distance between **X**^(*d*)^ and **Y**^(*d*)^ as defined in Equation 1. Similar to the one-dimensional DTW distance, we could also introduce the Sakoe-Chiba constraint on each dimension of *DTW*_*I*_. For the joint diet and physical activity time series in this paper, the two dimensions are largely different in scales and units. Also, we wish to study the influences on the clusters we generate as we change the emphasis on physical activity and diet. Therefore, we introduce a parameter *α* to bring the diet dimension and the physical activity dimension of the joint time series into similar scales and also to control the emphasis on physical activity and diet. In this paper, the independent multi-dimensional DTW with Sakoe-Chiba Band (*CDTW*_*I*_) for the joint diet and physical activity time series is defined as:

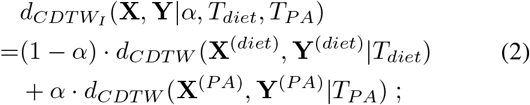

where **X** and **Y** are two joint diet and physical activity time series, and **X**^(*diet*)^*/***Y**^(*diet*)^ and **X**^(*PA*)^*/***Y**^(*PA*)^ are their diet dimensions and physical activity dimensions. *d*_*CDT W*_ (·) is the one-dimensional Constrained DTW with the Sakoe-Chiba Band. *T*_*diet*_ and *T*_*PA*_ are the Sakoe-Chiba Bandwidth for diet and physical activity dimensions respectively. *α* is the parameter that controls the emphasis on physical activity over diet (0 ≤ *α* ≤ 1). Larger *α* indicates that participants’ physical activity difference would have greater influence on their *CDTW*_*I*_ distance. When *α* = 1, the *CDTW*_*I*_ distance is equivalent to one-dimensional DTW computed only using the physical activity data. Analogously, the *CDTW*_*I*_ distance is equivalent to one-dimensional DTW computed only using diet data when *α* = 0.

Compared to *DTW*_*I*_, the dependent multi-dimensional DTW with Sako-Chiba Band (denoted as *CDTW*_*D*_) is defined in a similar way as the one-dimensional DTW in Equation 1:

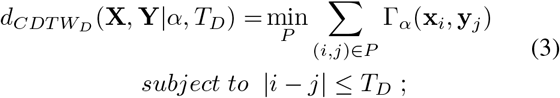

Compared to Equation 1, the samples of the joint time series in Equation 3 are *D*-dimensional vectors. Therefore, the corresponding local distance function needs to be Γ_*α*_ : ℝ^*D*^ × ℝ^*D*^ → ℝ^+^. Similar to *CDTW*_*I*_, we also introduce a parameter *α* in *CDTW*_*D*_ to control the scale of diet and physical activity and the emphasis on physical activity over diet. In *CDTW*_*D*_, *α* is included in the local distance function Γ_*α*_:

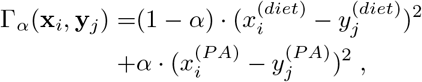

where 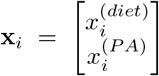 is the *i*^*th*^ sample of time series **X** (1 ≤ *i* ≤ *M*). 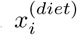 and 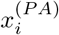 are the energy intake and Physical Activity Count (PAC) at the *i*^*th*^ minute of the day. For most values of *α*, the distances computed by *CDTW*_*D*_ and *CDTW*_*I*_ are different. When *α* = 1, they both converge to one-dimensional CDTW based only on the physical activity data, when *α* = 0, they both converge to one-dimensional CDTW based only on the diet data. Note that all dimensions in *CDTW*_*D*_ share the same Sakoe-Chiba Bandwidth *T*_*D*_, whereas each dimension in *CDTW*_*I*_ has its own bandwidth.

### D. Clustering Methods

After computing the multi-dimensional DTW distances using the joint diet and physical activity time series, the next step is to separate the participants into mutually exclusive clusters. In this paper, three distance-based clustering methods which are commonly used in time series clustering are combined with the multi-dimensional DTW distances, namely kernel k-means (KKM) [19], [47], spectral clustering (SPEC) [41], and kenerl hierarchical agglomerative clustering (KHAC) [31], [45]. In kernel hierarchical agglomerative clustering, the distance between two clusters is denoted as the linkage method. We explore four different ways of defining the linkage, including Single, Complete, Average, and Ward’s linkage. Our experiments show that only kernel k-means and KHAC with Ward’s linkage can generate more equal-sized clusters. As for clusters generated by the other clustering methods, there are usually one cluster that consists of over 90% of the entire dataset, leaving few participants in the other clusters. According to the NHANES Analytic Guidelines [4], a cluster size less than 30 is considered insufficient for inferential analysis based on normal approximation. In the sequel, we disregard the results generated by spectral clustering and KHAC with Single, Complete, and Average linkage, and focus on the most successful approaches for producing more equal-sized clusters including kernel k-means and KHAC with Ward’s linkage.

In this paper, the Gaussian Dynamic Time Warping Kernel [12] is used to convert *CDTW*_*D*_ and *CDTW*_*I*_ into kernel functions to combine with the distance-based clustering methods:

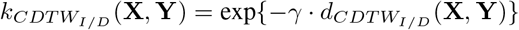

where *γ* is fixed to be half of the average of all pairwise distances. Due to the time complexity of multi-dimensional DTW, it takes a large amount of time to compute the pairwise distances of all participants using CPU. To address the computational issue, we exploit the parallel structure of graphic processing units (GPU) to accelerate the computation.

## IV. Experiments and Evaluation

### A. Dataset

The National Health and Nutrition Examination Survey (NHANES) is a cross-sectional survey designed to assess the nutritional and health status of the U.S. non-institutionalized civilian population [5]–[7]. The participants of NHANES were recruited through a complex, multi-stage probability sampling design. The NHANES is unique in that it is one of the few publicly-available and nationally representative datasets which combines physical examinations and interviews for data collection. The joint diet and physical activity dataset used in this paper is constructed from the NHANES dataset. In this paper, we exclude the participants who were pregnant, younger than 20, older than 65, or missing dietary, physical activity, anthropometric or laboratory data.

#### 1) Diet Data

The NHANES collected two days of dietary recalls from the participants using the USDA Automated Multiple-Pass Method [8]. The participants reported their food consumption during a 24-hour period for each recall, including the type and amount of each food, the time of intake, and food descriptions. All reported food consumption was converted into energy intake (kcal) according to the USDA Food and Nutrient Database for Dietary Studies (FNDDS) for 2003-2004 data [2] and 2005-2006 data [3]. In this paper, we focus on the participants’ weekday energy intakes for dietary assessment, and participants whose dietary recalls are both from weekend were excluded. From the two dietary recalls of the remaining participants, the first one is selected to form the dietary dataset if it is a weekday, otherwise, the second recall is selected. The original energy intakes were reported with a time stamp (minute-level) indicating when the eating occasions started, but the duration of the eating occasions is not collected by the NHANES. To approximate real-life eating situations in the diet time series, we assume that each eating occasion takes 15 minutes based on a previous study [35], and the energy intakes were smoothed by an average filter of length 15 (minute) to generate the final diet time series.

#### 2) Physical Activity Data

The physical activity data were collected by uni-axial accelerometers. The NHANES required the participants to wear an ActiGraph AM-7164 [5] on the right hip. The Actigraph AM-7164 measures acceleration in the vertical direction in units knwon as “Physical Activity Count (PAC)”. The original accelerometer measurements (10*Hz* sampling frequency) were filtered and calibrated by the devices to achieve linear associations between PACs and a measured physiologic variable [29]. In the NHANES 2003-2006 Examination, the PACs (10*Hz*) were further summed over each one-minute epoch [5]. To summarize, the physical activity time series in the NHANES are minute-level PACs filtered and digitized to reflect activity intensity [27]. All participants were required to keep the devices on for 7 consecutive days. Due to compliance and other factors, there are a large number of participants who do not have a full 7-day record. We focus on the participants’ weekday physical activity patterns. To maximize the number of participants involved in this study, we include anyone with at least one weekday of valid accelerometer data. For those participants with multiple valid weekday data, one valid weekday is randomly selected to form the daily physical activity dataset.

#### 3) Joint Diet and Physical Activity Data

The participants in the joint diet and physical activity dataset are the intersection of the diet dataset and the physical activity dataset described above. There are 1836 targeted participants after exclusions. We further combine these participants’ diet and physical activity data into two-dimensional joint time series. The samples of the joint diet and physical activity time series are two-dimensional vectors of energy intakes and PACs, i.e., the joint diet and physical activity time series has the following format:

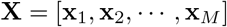

where 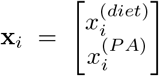 is the *i*^*th*^ sample of time series **X** (1 ≤ *i* ≤ *M, M* = 1440). 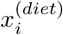 and 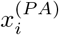 represent the energy intake and PAC at the *i*^*th*^ minute of the day.

The scales of the energy intakes and PACs can be largely different. Since we have introduced the parameter *α* as a multiplier when computing the multi-dimensional DTW distances, we do not globally standardize the diet and the physical activity time series (subtracting from global mean and dividing by global standard deviation) before combining them into joint time series. This is reasonable in that subtracting all the time series by the same mean has no influences on the computed DTW distances, and dividing by the standard deviation can be achieved by adjusting the value of *α*. In some time series studies [13], [17], [25], [55], it is common to use z-normalization [26] as a data pre-processing step. Mathematically, z-normalize a time series **X** is to subtract from its mean and divide by its deviation: 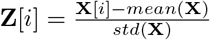. From our experiments, using z-normalization to pre-process the diet or the physical activity time series weakens the influences of different amount of energy intake or intensity of physical activity. In the sequel, the clusters derived with z-normalization have little difference in terms of health. Therefore, we do not include the clustering results based on z-normalized data in this paper.

### B. Cluster Evaluation

The joint diet and physical activity time series are clustered using methodologies discussed in Section III. For both *CDTW*_*D*_ and *CDTW*_*I*_ distances, parameter *α* controls the emphasis on physical activity over diet. Parameter *T*_*D*_ is the Sakoe-Chiba Bandwidth which constrains the maximum time difference between aligned samples in *CDTW*_*D*_. Analogously, parameter *T*_*diet*_ and *T*_*PA*_ are the Sakoe-Chiba Band-widths in *CDTW*_*I*_, each of which constrains a single dimension of diet or physical activity. With different values of *α* and the Sakoe-Chiba Bandwidths, the multi-dimensional DTW distances would have varying emphasis on physical activity over diet (controlled by *α*), or temporality over intensity (controlled by *T*). Theoretically, the Sakoe-Chiba Bandwidth can be any integer value from 0 to 1440, and *α* can be any value from 0 to 1. Due to computational limitations, we only investigate a limited number of values for each parameter. These parameter values are selected such that the visualizations corresponding to different parameter values have noticeable differences. In this paper, we investigate 4 values for the Sakoe-Chiba Bandwidths ranging from 120 to 480 (minute) in steps of 120, and 21 values for *α* ranging from 0.000 to 0.040 in steps of 0.002, i.e., there are 84 (21 *α* × 4 *T*_*D*_) parameter combinations for *CDTW*_*D*_ and 336 (21 *α* × 4 *T*_*diet*_ × 4 *T*_*PA*_) for *CDTW*_*I*_. The values of *α* may appear to be rather small. This does not indicate that the DTW distances focus only on the diet and neglect the physical activity. As discussed in Section IV-A, since we do not standardize the diet and the physical activity data before combining them into joint time series, *α* is used to bring the diet and the physical activity data into similar scale. It can be seen in Figure 1 that the mean trajectories of the clusters appear to be different even for small values of *α*. The multi-dimensional DTW with different parameter combinations can be seen as different distance measures, and we combine each distance measure with all the clustering methods mentioned in Section III-D. In the following discussion, we will focus on kernel k-means and KHAC with Ward’s linkage as they are most successful in producing more equal-sized clusters. To find a proper number of clusters, we test four values (*k* ∈ {3, 4, 5, 6}) under each combination of distance measure and clustering method.

**Fig. 1.**
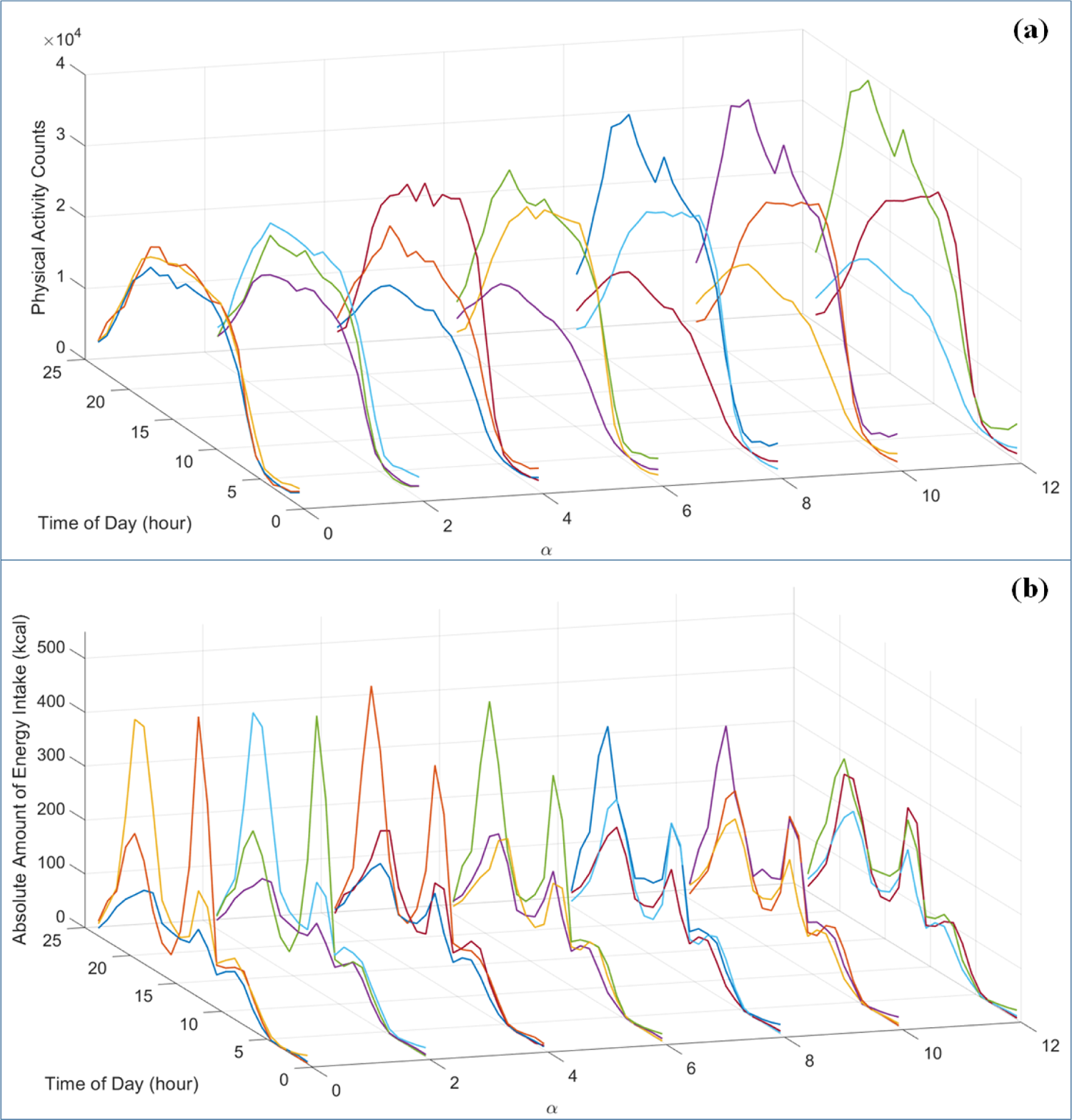
The visualizations of the clustering results corresponding to different values of *α*, where the clustering method and distance measure are fixed to kernel k-means and *CDTW*_*D*_, the Sakoe-Chiba Bandwidth of *CDTW*_*D*_ is fixed to *T*_*D*_ = 120. At each *α*, the YZ-planes in (a) and (b) are the physical activity visualization and the diet visualization of the same clustering result.

From all the combinations of distance measures and clustering methods we explore, we wish to select the ones that could generate temporal joint diet and physical activity patterns (TJDPAP) which have distinctive physical activity and diet characteristics, as well as meaningful links to health. Following our previous temporal pattern studies [22], [24], [33], we use three approaches for evaluating the clustering results in this paper, including the visualizations of the clusters, the Silhouette Index (internal criteria), and the associations between clusters and health status indicators determined by multivariate regression models (external criteria).

#### 1) Cluster Visualization

here are various ways to visualize a cluster of time series such as mean trajectory, heat map, and DTW Barycenter Averaging [43]. We focus on the mean trajectories for cluster visualization as they are most intuitive for showing the traits of the clusters. For a cluster **C** of joint diet and physical activity time series **X**_*i*_, its mean trajectories corresponding to the diet dimension and the physical activity dimension are defined as:

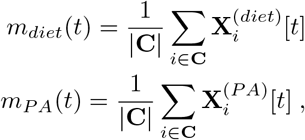

where |**C**| is the number of joint time series in cluster **C**. 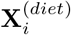 and 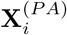 are the diet dimension and the physical activity dimension of the joint time series **X**_*i*_. The time unit is converted into hour-level for better visualization, i.e., 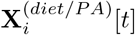 is the summed PACs or energy intakes over the *t*^*th*^ hour in time series **X**_*i*_ (*t* ∈ [0 : 23]).

Figure 1 shows the visualizations of the clustering results corresponding to different values of *α*, where the clustering method and distance measure are fixed to kernel k-means and *CDTW*_*D*_, the Sakoe-Chiba Bandwidth of *CDTW*_*D*_ is fixed to *T*_*D*_ = 120. Figure 1 (a) and (b) are the mean trajectories of the physical activity dimension and the diet dimension respectively. In Figure 1, the values of *α* are from 0.000 to 0.012 in steps of 0.002. We did not include the rest of the values for *α* because there are no significant changes in the clustering results after *α >* 0.012. At each *α*, the YZ-planes in Figure 1 (a) and (b) are the physical activity visualization and the diet visualization of the same clustering result. As the value of *α* increases, we focus more on the physical activity over diet while computing the *CDTW*_*D*_ distances, and this change of focus is reflected in Figure 1: When *α* is small (e.g., *α* = 0), the diet differences between the clusters are most significant and the mean trajectories of the physical activity are very similar; when *α* is large (e.g., *α* ≥ 0.012), the physical activity differences become more dominant.

#### 2) Internal Criteria

The internal criteria defines the goal of achieving higher intra-cluster similarity and lower inter-cluster similarity [10]. When ground truth is not available, the number of clusters is commonly determined by the internal criteria [48], [59]. Following this practice, we use the Silhouette Index [46] to find the number of clusters. We test four values *k* ∈ {3, 4, 5, 6} under each combination of clustering method and distance measure. From our experiments, the choice of *k* = 3 generally achieved higher Silhouette Index in most situations, indicating a better clustering quality when the number of clusters equals 3 (due to page limitations, we did not list the values of the Silhouette Index in this paper). Therefore, we select the number of clusters to be *k* = 3 in the following discussion.

#### 3) External Criteria

The external criteria evaluates the clustering results based on a priori information. In the NHANES dataset, each participant has several health status indicators such as body mass index (BMI) and blood pressure. Following the practice in the previous temporal pattern studies [11], [22], [23], [32], we use the health status indicators to define an external criterion which evaluates the clusters’ association to health. For each health status indicator, we perform Multivariate Linear Regression (MLR) and multiple comparison analysis with the cluster labels as explanatory variable, and two clusters are significantly different regarding this health status indicator if their adjusted p-value from multiple comparison is less than the 5% significance level. Twelve health status indicators are included to define the external criterion because of their previous associations with physical activity or diet [16], [20], [56], and the external criterion is the total number of pairwise cluster comparisons that are significantly different. With more pairs of clusters showing significant differences in health status indicators, there is a stronger association between the clusters and health. Therefore, larger values of the external criteria indicate better clustering performance. For detailed information regarding the selected health status indicators, we refer readers to the Anthropometric Assessment and Laboratory Tests section in [22], [32].

Figure 2 and Figure 3 are the external criteria for the clustering derived using *CDTW*_*I*_ combined with kernel k-means (KKM) and kernel hierarchical agglomerative clustering (KHAC) respectively. In Figure 2 and Figure 3, the Y-axis represents the value of the external criteria, and the points are the clusters derived from different parameter combinations. To determine the parameter combinations of a specific clustering, parameter *α* can be found from its X-axis, parameter *T*_*PA*_ (the Sakoe-Chiba Bandwidth for physical activity dimension) can be found by the color, and parameter *T*_*Diet*_ (the Sakoe-Chiba Bandwidth for diet dimension) can be found by the symbol. Similarly, Figure 4 shows the results derived using *CDTW*_*D*_ combined with kernel k-means and kernel hierarchical agglomerative clustering. In Figure 4, the clustering method of a point (clustering result) can be found by the color, and parameter *T*_*D*_ can be found by the symbol.

**Fig. 2.**
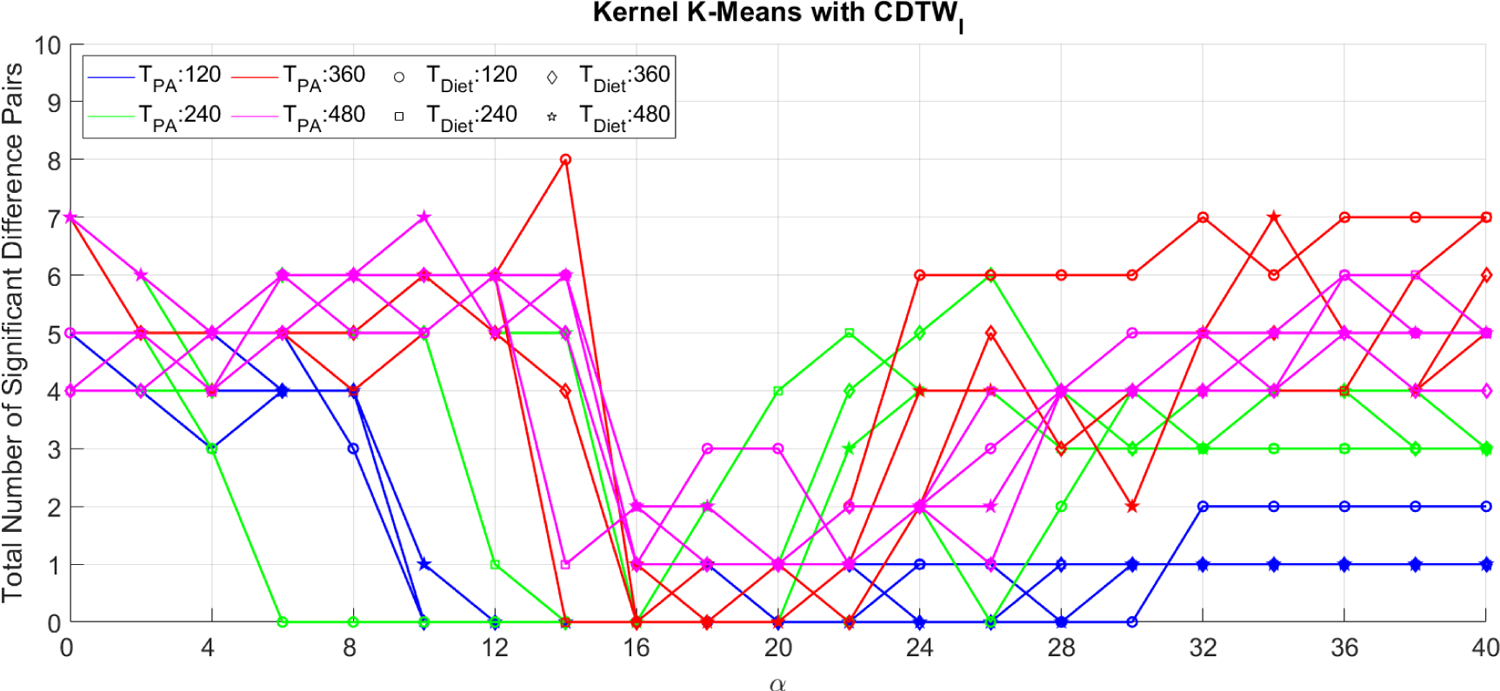
The external criteria for clustering results derived using kernel k-means and *CDTW*_*I*_.

**Fig. 3.**
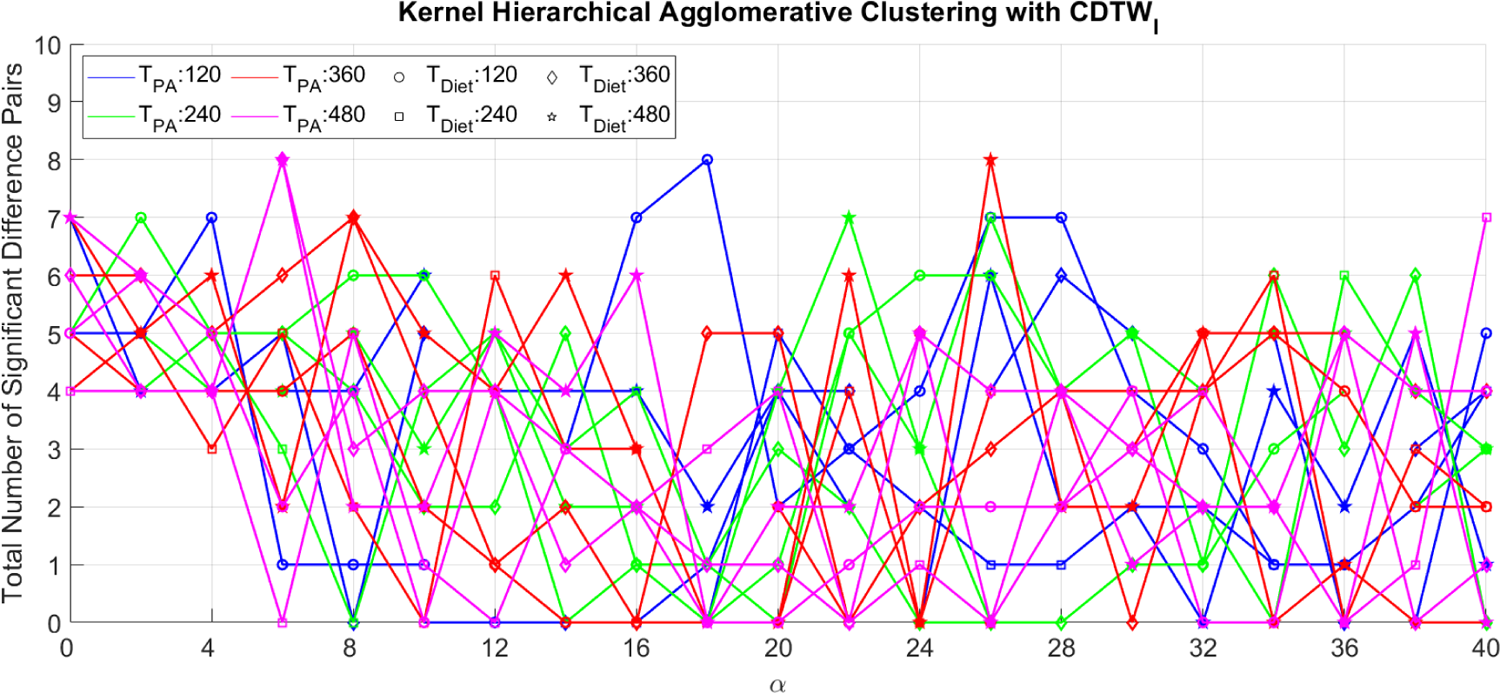
The external criteria for clustering results derived using kernel hierarchical agglomerative clustering and *CDTW*_*I*_.

**Fig. 4.**
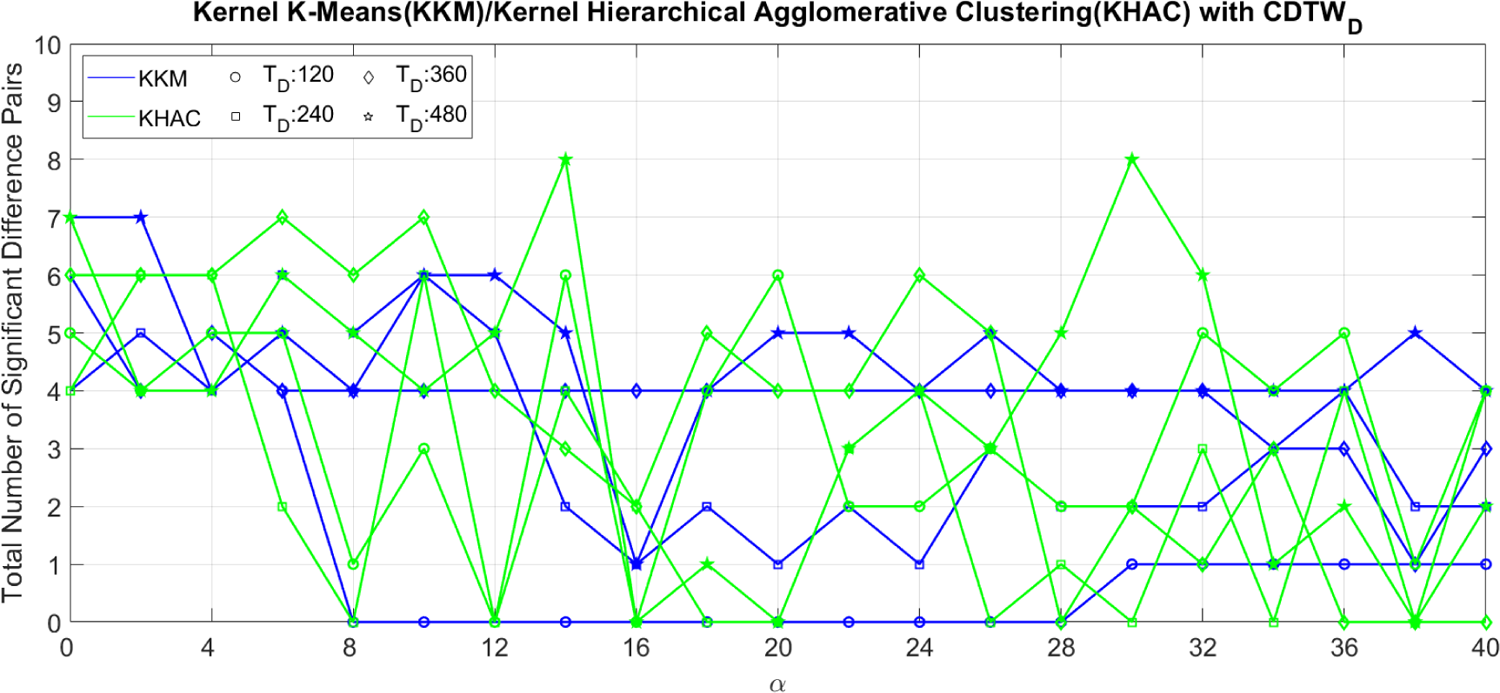
The external criteria for clustering results derived using kernel hierarchical agglomerative clustering/kernel k-means and *CDTW*_*D*_.

From Figure 2, Figure 3, and Figure 4, there are seven clustering results that achieve the highest external criteria (8 pairs of significantly different clusters among all health status indicators): KHAC combined with *CDTW*_*I*_ at (*T*_*PA*_ = 120, *T*_*Diet*_ = 120, *α* = 0.018), (*T*_*PA*_ = 360, *T*_*Diet*_ = 480, *α* = 0.026), (*T*_*PA*_ = 480, *T*_*Diet*_ = 360, *α* = 0.006), and (*T*_*PA*_ = 480, *T*_*Diet*_ = 480, *α* = 0.006); KHAC combined with *CDTW*_*D*_ at (*T*_*D*_ = 480, *α* = 0.014) and (*T*_*D*_ = 480, *α* = 0.030); and KKM combined with *CDTW*_*I*_ at (*T*_*PA*_ = 360, *T*_*Diet*_ = 120, *α* = 0.014). Compared with KKM, KHAC is more likely to generate clustering results which are more related to health. However, KHAC is more sensitive to small changes in the distance matrix. This can be shown in Figure 3, and Figure 4 where the curves of the KHAC have more fluctuations compared with KKM. In terms of the two distance measures, *CDTW*_*I*_ has the flexibility to combine different constraints on the diet and physical activity dimension, thus, it has the advantage to generate clustering results with more varieties and stronger links to health. This is partially due to the fact that the diet and the physical activity data are loosely coupled and do not follow the same routine in a day.

## V. Conclusion

In this paper, we described a distance-based cluster analysis approach to find joint temporal diet and physical activity patterns among U.S. adults. Two multi-dimensional DTW distances, *CDTW*_*I*_ and *CDTW*_*D*_, are combined with kernel k-means, kernel hierarchical agglomerative clustering with Ward’s linkage, and several other clustering methods to generate the joint patterns. The clustering results are evaluated using visualization of the clusters, the Silhouette Index, and the associations between clusters and health status indicators based on multivariate regression models.

From the visualizations of the clusters, the parameters of multi-dimensional DTW distances give us the flexibility to control the clustering focus on physical activity over diet (controlled by parameter *α*), and on temporality over intensity (controlled by the Sakoe-Chiba Bandwidths). Diet patterns and physical activity patterns seem to have weak correlation as the mean trajectories of physical activity when *α* = 0 and the mean trajectories of diet when *α* = 1 are not distinguishable. From our experiments, most clustering results that have the stronger associations to health (largest number of significant difference pairs) are generated with 0.006 ≤*α* ≤0.030. As shown in Figure 1, these values of *α* indicate that the multi-dimensional DTW distances have a mixed focus on both diet and physical activity compared to *α* = 0 (only diet is considered) and *α* = 1 (only physical activity is considered). This demonstrates that the integration of diet, physical activity, and time, has the potential to find joint temporal patterns with stronger associations to health.

## Data Availability

the National Health and Nutrition Examination Survey website

https://wwwn.cdc.gov/nchs/nhanes/ContinuousNhanes/Default.aspx?BeginYear=2003

